# Two-stage biologically interpretable neural-network models for liver cancer prognosis prediction using histopathology and transcriptomic data

**DOI:** 10.1101/2020.01.25.20016832

**Authors:** Zhucheng Zhan, Zheng Jing, Bing He, Noshad Hosseini, Maria Westerhoff, Eun-Young Choi, Lana X. Garmire

## Abstract

**Purpose:** Pathological images are easily accessible data with the potential as prognostic biomarkers. Moreover, integration of heterogeneous data types from multi-modality, such as pathological image and gene expression data, is invaluable to help predicting cancer patient survival. However, the analytical challenges are significant.

**Experimental Design:** Here we take the hepatocellular carcinoma (HCC) pathological image features extracted by CellProfiler, and apply them as the input for Cox-nnet, a neural network-based prognosis. We compare this model with conventional Cox-PH model, CoxBoost, Random Survival Forests and DeepSurv, using C-index and log ranked p-values on HCC testing samples. Further, to integrate pathological image and gene expression data of the same patients, we innovatively construct a two-stage Cox-nnet model, and compare it with another complex neural network model PAGE-Net.

**Results:** pathological image based prognosis prediction using Cox-nnet is significantly more accurate than Cox-PH and random survival forests models, and comparable with CoxBoost and DeepSurv methods. Additionally, the two-stage Cox-nnet complex model combining histopathology image and transcriptomics RNA-Seq data achieves better prognosis prediction, with a median C-index of 0.75 and log-rank p-value of 6e-7 in the testing datasets. The results are much more accurate than PAGE-Net, a CNN based complex model (median C-index of 0.68 and log-rank p-value of 0.03). Imaging features present additional predictive information to gene expression features, as the combined model is much more accurate than the model with gene expression alone (median C-index 0.70). Pathological image features are modestly correlated with gene expression. Genes having correlations to top imaging features have known associations with HCC patient survival and morphogenesis of liver tissue.

**Conclusion:** This work provides two-stage Cox-nnet, a new class of biologically relevant and relatively interpretable models, to integrate multi-modal and multiple types of data for survival prediction.

## Introduction

Prognosis prediction is important for providing effective disease monitoring and management. Various biomaterials have been proposed as potential biomarkers to predict patient survival. Among them, hematoxylin and eosin (H&E) stained histopathological images, are very attractive materials to extract biomarker features. Compared to genomics materials, such as RNA-Seq transcriptomics, these images are much more easily accessible and cheaper to obtain, through processing archived formalin-fixed paraffin-embedded (FFPE) Blocks. In H&E staining, the hematoxylin is oxidized into phematein, a basic dye which stains acidic (basophilic) tissue components (ribosomes, nuclei, and rough endoplasmic reticulum) into darker purple color. Whereas acidic eosin dye stains other protein structures of the tissue (stroma, cytoplasm, muscle fibers) into a pink color. As patients’ survival information is retrospectively available in electronic medical record data and FFPE blocks are routinely collected clinically, the histopathology images can be generated and used for highly valuable and predictive prognosis models.

Previously, we developed a neural network model called Cox-nnet to predict patient survival, using transcriptomics data (1). Cox-nnet is an alternative to the conventional methods, such as Cox proportional hazards (Cox-PH) methods with LASSO or ridge penalization. We have demonstrated that Cox-nnet is more optimized for survival prediction from high throughput gene expression data, with comparable or better performance than other conventional methods, including Cox-PH, Random Survival Forests (2) and Coxboost (3). Moreover, Cox-nnet reveals much richer biological information, at both the pathway and gene levels, through analysing the survival related “surrogate features’’ represented as the hidden layer nodes in Cox-nnet. A few other neural network based models were also proposed around the same time as Cox-nnet, such as DeepSurv (4). It remains to be explored if Cox-nnet can take input features from other data types that are less biologically intuitive than genomics data, such as histopathology imaging data. It is also important to benchmark Cox-nnet with the other above mentioned methods. Moreover, some neural network based models were reported to handle multi-modal data (5). For example, PAGE-Net is a complex neural network model that has a convolutional neural network (CNN) layer followed by pooling and a genomics model involved in transformation of the gene layer to pathway layer (5). The genomics neural network portion is followed by two hidden layers, the latter of which is combined with the image neural network model to predict glioblastoma patient survival. Though PAGE-Net uses CNN, the resulting predictive C-index value based on imaging data raises concern of overfitting (train C-index=0.97; test C-index=0.68). It is therefore important to test if a model built upon Cox-nnet, using pre-extracted, biologically informative features, can combine multiple types of data, eg. imaging and genomics data, and if so, how well it performs relative to models such as PAGE-Net.

In this study, we extend Cox-nnet to take up pathological image features extracted from imaging processing tool *CellProfiler* (6), and compare the predictive performance of Cox-nnet relative to Cox proportional hazards, the standard method for survival analysis, which was also the second best method in the previous survival prediction study using pan-cancer datasets (1). We further used the pathology image data to compare Cox-nnet with two other methods CoxBoost and Random Survival Forests, which we had compared before on genomics data, as well as the start of the art method DeepSurv (4). Moreover, we propose a new type of two-stage complex Cox-nnet model, which combines the hidden node features from multiple first-stage Cox-nnet models, and then use these combined features as the input nodes to train a second stage Cox-nnet model. We applied the models on TCGA hepatocellular carcinoma (HCC), which we had previously gained domain experience on (7,8). Hepatocellular carcinoma (HCC) is the most prevalent type of liver cancer that accounts for 70%-90% of all liver cancer cases. It is a devastating disease with poor prognosis, where the 5-year survival rate is only 12% (9). And the prognosis prediction becomes very challenging due to the high level of heterogeneity in HCC as well as the complex etiologic factors. Limited treatment strategies in HCC, relative to other cancers, also imposes an urgent need to develop tools for patient survival prediction. As comparison, we also evaluated the performance of another CNN based model called PAGE-Net, and showed that Cox-nnet achieves higher accuracy in testing data.

## Material & Methods

### Datasets

The histopathology images and their associated clinical information are downloaded from The Cancer Genome Atlas (TCGA). A total of 384 liver tumor images are collected. Among them 322 samples are clearly identified with tumor regions by pathology inspection. Among these samples, 290 have gene expression RNA-Seq data, and thus are selected for pathology-gene expression integrated prognosis prediction. The gene expression RNA-Seq dataset is also downloaded from TCGA, each feature was then normalized into RPKM using the function *ProcessRNASeqData* by TCGA-Assembler (10).

### Tumor Image Pre-processing

For each FFPE image stained with H&E, two pathologists at University of Michigan provide us with the ROI (tumor regions). The tumor regions are then extracted using Aperio software *ImageScope* (11). To reduce computational complexities, each extracted tumor region is divided into non-overlapping 1000 by 1000 pixel tiles. The density of each tile is computed as the summation of red, green and blue values, and 10 tiles with the highest density are selected for further feature extraction, following the guideline of others (12). To ensure that the quantitative features are measured under the same scale, the red, green and blue values are rescaled for each image. Image #128 with the standard background color (patient barcode: TCGA-DD-A73D) is selected as the reference image for the others to be compared with. The means of red, green and blue values of the reference image are computed and the rest of the images are normalized by the scaling factors of the means of red, green, blue values relative to those of the reference image.

### Feature extraction from the images

CellProfiler is used for feature extraction (13). Images are first preprocessed by ‘*UnmixColors*’ module to H&E stains for further analysis. ‘*IdentifyPrimaryObjec*t’ module is used to detect unrelated tissue folds and then removed by ‘*MaskImage*’ module to increase the accuracy for detection of tumour cells. Nuclei of tumour cells are then identified by *‘IdentifyPrimaryObject’* module again with parameters set by Otsu algorithm. The identified nuclei objects are utilised by ‘*IdentifySecondaryObject*’ module to detect the cell body objects and cytoplasm objects which surround the nuclei. Related biological features are computed from the detected objects, by a series of feature extraction modules, including *‘MeasureGranularity’, ‘MeasureObjectSizeShape’, ‘MeasureObjectIntensity’, ‘MeasureObjectIntensityDistribution’, ‘MeasureTexture’, ‘MeaureImageAreaOccupied’, ‘MeasureCorrelation’, ‘MeasureImageIntensity’* and *‘MeasureObjectNeighbors*’. To aggregate the features from the primary and secondary objects, the related summary statistics (mean, median, standard deviation and quartiles) are then calculated to summarize data from object level to image level, yielding 2429 features in total. Each patient is represented by 10 images, and the median of each feature is selected to represent the patient’s image biological feature.

### Survival prediction models

#### Cox-nnet model

The Cox-nnet model is implemented in the Python package named Cox-nnet (1). Current implementation of Cox-nnet is a fully connected, two-layer neural network model, with a hidden layer and an output layer for cox regression. A drop-out rate of 0.7 is applied to the hidden layer to avoid overfitting. The size of the hidden layer is equal to the square root of the number of input features. We used a hold-out method by randomly splitting the dataset to 80% training set and 20% testing set, using *train_test_split* function from sklearn package. We used grid search and 5-fold cross-validation to optimise the hyper-parameters for the deep learning model on the selected training set. We trained the model with a learning rate of 0.01 for 500 epochs with no mini-batch applied, then evaluated the model on testing data. The procedure is repeated 20 times to assess the average performance. More details about Cox-nnet is described earlier in Ching et al (1).

#### Cox proportional hazards Model

Since the number of features produced by *CellProfiler* exceeds the sample size, an elastic net Cox proportional hazard model is built to select features and compute the prognosis index (PI) (14). Function *cv*.*glmnet* in the *Glmnet* R package is used to perform cross-validation to select the tuning parameter *lambda*. The parameter *alpha* that controls the trade-off between quadratic penalty and linear penalty is selected using grid search. Same hold-out setting is employed by training the model using 80% randomly selected data and evaluated on the remaining 20% testing set. The procedure is repeated 20 times to calculate the mean accuracy of the model.

#### CoxBoost Model

CoxBoost is a modified version of Cox-PH model, but is especially suited for models with a large number of predictors (15). As an iterative gradient boosting method, CoxBoost separates the parameters into individual partitions. It first selects the partition that leads to the largest improvement in the penalized partial log likelihood, then selects other blocks in subsequent iterations and refits the parameters by maximizing the penalized partial log likelihood.

#### Random Survival Forest Model

Random Survival Forests is a non-linear tree-based ensemble method (16). Each tree in the forest is fitted on bootstrapped data, with nodes split by maximizing the log-rank statistics. A patient’s cumulative hazard is then calculated as the averaged cumulated hazard over all trees in the ensemble. We implemented RFS using R package ‘randomForestSRC’ (17), where the hyperparameters such as number of trees or node size are optimized by random search in R.

#### DeepSurv Model

DeepSurv is a deep learning generalization of the Cox proportional hazards model, for predicting a patient’s risk of failure (4). DeepSurv is implemented in Python using Theano and Lasagne, with hyperparameters optimized by random search over box constraints. The optimal DeepSurv architecture for our imaging data was determined by its built-in random hyperparameter search, consisting of two hidden layers with 38 nodes and 0.07 dropout for each. We trained DeepSurv model on image data with a learning rate of 1e-5 and a learning rate decay of 6e-4 for 10,000 epochs.

#### Two-stage Cox-nnet model

The two-stage Cox-nnet model has two phases, as indicated in the name. For the first stage, we construct two separate Cox-nnet models in parallel, one for the image data and the other one for gene expression data. For each model, we optimize the hyper-parameters using grid search under 5-fold cross-validation, as described earlier. In the second stage, we extract and combine the nodes of the hidden layer from each Cox-nnet model as the new input features for a new Cox-nnet model. We construct and evaluate the second stage Cox-nnet model with the same parameter-optimisation strategy as in the first-stage.

#### PAGE-Net model

it is another neural-network method that can combine imaging and genomics (eg. gene expression) information to predict patient survival (5). The imaging prediction module is very complex, with a patch-wide pre-trained convolutional neural network (CNN) layer followed by pooling them together for another neural network. The pre-trained CNN of PAGE-Net consists of an input layer, three pairs of dilated convolutional layers (a kernel size of 5*5, 50 feature maps, and a dilation rate of 2) and a max-pooling layer of 2*2 size. The sequential layers are followed by a flatten layer and a fully connected layer. The size of patches for training CNN is 256*256. After CNN, PAGE-Net consists of one gene layer (4675 nodes), one pathway layer (659 nodes), two hidden layers (100 nodes; 30 nodes), one pathology layer (30 nodes) and a cox output layer. Drop-out rates of 0.7, 0.5, 0.3 are applied to two hidden layers and pathology hidden layer respectively. We trained PAGE-Net with a learning rate of 1.5e-6 and a batch size of 32 for 10,000 epochs. The hyperparameters are optimized by grid search. It’s noteworthy that PAGE-Net has serious running time issues in CNN pretraining and feature pooling steps, therefore we only repeated the integrative layer training with 20 different train-test splits.

### Model evaluation

Similar to the previous studies (1,7,8), we also use concordant index (C-index) and log-rank p-value as the metrics to evaluate model accuracy. C-index signifies the fraction of all pairs of individuals whose predicted survival times are correctly ordered and is based on Harrell C statistics. The equation is as follows:

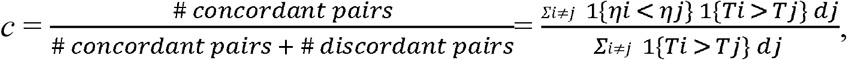

where *η* is the predicted risk score, *T* is the “time-to-event” response, *d* is an auxiliary variable such that *d*=1 if event is observed and *d*=0 if patient is censored. A C-index of 1 means the model fits the survival data perfectly, whereas a score around 0.50 means randomness. In practice, a C-index around 0.70 indicates a good model. As both Cox-nnet and Cox-PH models quantify the patient’s prognosis by log hazard ratios, we use the predicted median hazard ratios to stratify patients into two risk groups (high vs. low survival risk groups). We also compute the log-rank p-value to test if two Kaplan-Meier survival curves produced by the dichotomised patients are significantly different, similar to earlier reports (7,14,18-21).

### Feature evaluation

The input feature importance score is calculated by drop-out. The values of a variable are set to its mean and the log likelihood of the model is recalculated. The difference between the original log likelihood and the new log likelihood is considered as feature importance (22). We select 100 features with the highest feature scores from Cox-nnet for association analysis between pathology image and gene expression features. We regress each of the top 10 image features (y) on each of the top 50 gene expression features (x), and use the R-square statistic with significant p-value (<0.05) as the correlation metric.

### Code availability

The code for two-stage Cox-nnet, including integration of hidden nodes, feature extraction, and feature analysis are all available at github: https://github.com/lanagarmire/two-stage-cox-nnet

## Results

### Overview of Cox-nnet model on pathological image data

In this study, we test if pathological images can be used to predict cancer patients. The initial task is to extract image features that can be used as the input for the predictive models. As described in the Methods, pathological images of 322 TCGA HCC patients are individually annotated with tumor contents by pathologists, before being subject to a series of processing steps. The tumor regions of these images then undergo segmentation, and the top 10 tiles (as described in section 2.2) out of 1000 by 1000 tiles are used to represent each patient. These tiles are next normalized for RGB coloring against a common reference sample, and 2429 image features of different categories are extracted by *CellProfiler*. Summary statistics (mean, median, standard deviation and quartiles) are calculated for each image feature, and the median values of them over 10 tiles are used as the input imaging features for survival prediction.

We apply these imaging features on Cox-nnet, a neuron-network based prognosis prediction method previously developed by our group. The architecture of Cox-nnet is shown in **Figure 1**. Briefly, Cox-nnet is composed of the input layer, one fully connected hidden layer and an output “proportional hazards’’ layer. We use 5-fold cross-validation (CV) to find the optimal regularization parameters. Based on the results on RNA-Seq transcriptomics previously, we use dropout as the regularization method.

**Figure 1:**
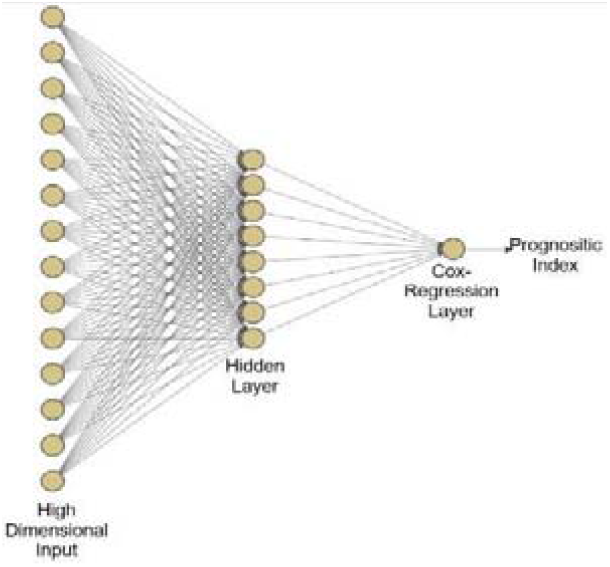
The architectures of Cox-nnet model: (The sketch of Cox-nnet model for prognosis prediction, based on a single data type.

### Comparison of prognosis prediction among Cox-nnet and other methods over pathology imaging data

To evaluate the results on pathology image data, we compare Cox-nnet with the three other models which were benchmarked before using genomics data: Cox-PH model, CoxBoost and Random Survival Forests (15), as well as another state of art neural network based method DeepSurv (**Figure 2**). We use two accuracy metrics to evaluate the performance of models in comparison: C-index and log-rank p-values. C-index measures the fraction of all pairs of individuals whose predicted survival times are correctly ordered by the model. The higher C-index, the more accurate the prognosis model is. As shown in **Figure 2A**, on the testing datasets, the median C-index score from Cox-nnet (0.74) is significantly higher (p<0.001) than both Cox-PH (0.72) and Random Survival Forests (0.70). Moreover, Cox-nnet achieves a comparable C-index score with other two models, CoxBoost (0.75, p=0.10) and DeepSurv (0.74, p=0.19). In particular, Cox-nnet yields more stable models than DeepSurv, as shown by the smaller variations in the C-index scores in different test sets. Additionally, we dichotomize survival risks using the median score of predicted prognosis index (PI) from each model. We then use log-rank p-value to show the survival difference between the Kaplan-Meier survival curves of high vs. low survival risk groups in a typical run(**Figure 2B-F** and **Supplementary Figure 3A-E**). In the training dataset Cox-nnet achieves the third highest log-rank p-value of 1e-12 (**Supplementary Figure 3A**), after those of DeepSurv (3e-19) and RSF (2e-21). Whereas in the testing dataset, Cox-nnet achieves the highest log-rank p-value of 4e-6, better than those of Cox-PH (3e-4), DeepSurv (7e-6), CoxBoost (3e-4), and Random Survival Forests (0.004). We next investigate the top 100 image features according to Cox-nnet ranking (**Supplementary Figure 1**). Interestingly, the most frequent features are those involved in textures of the image, accounting for 48% of raw input features. Intensity and Area/Shape parameters make up the 2nd and 3rd highest categories, with 18% and 15% features. Density, on the other hand, is less important (3%). It is also worth noting that among the 47 features selected by the conventional Cox-PH model, 70% (33) are also found in the top 100 features selected by Cox-nnet, showing the connections between the two models.

**Figure 2:**
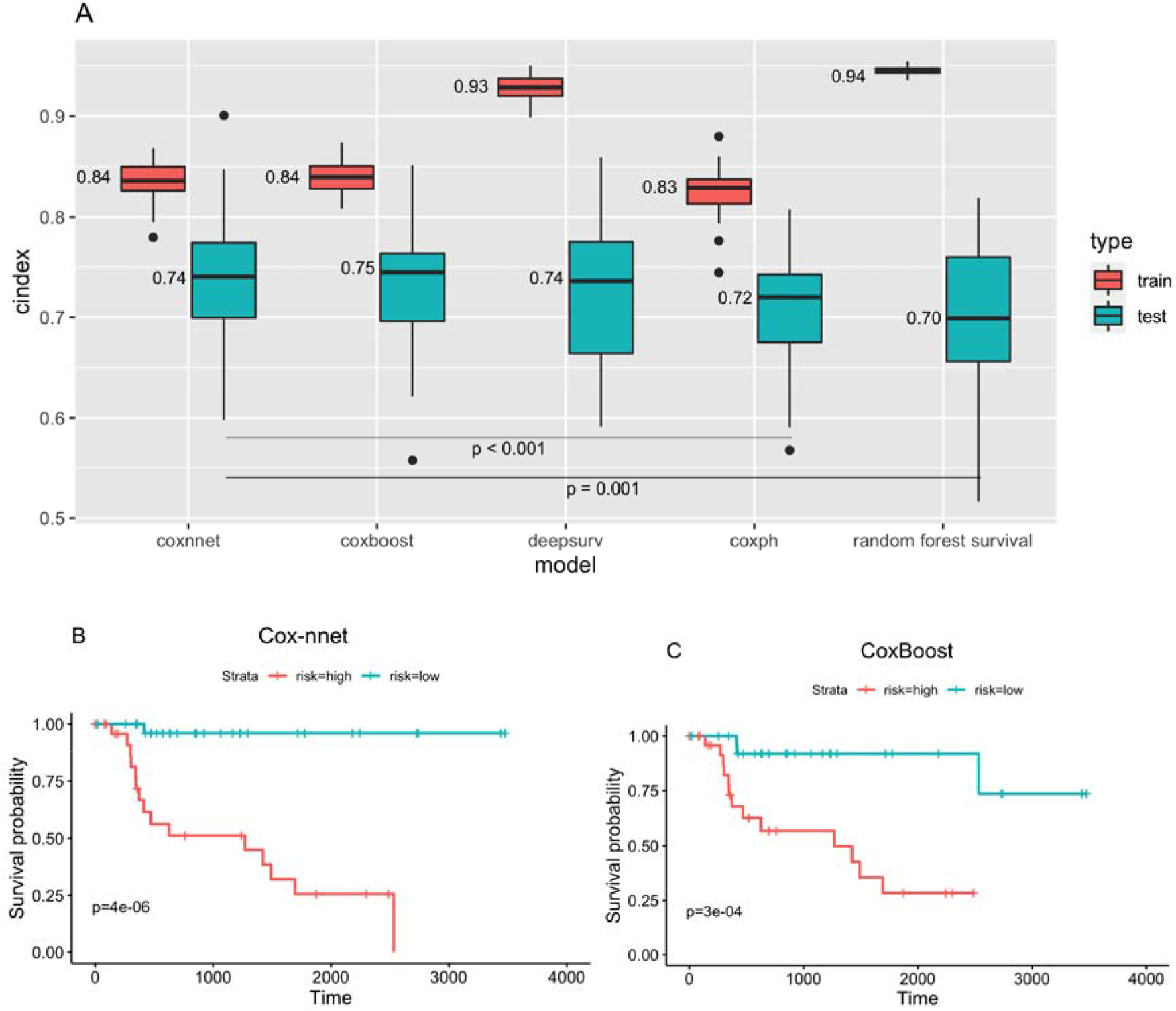
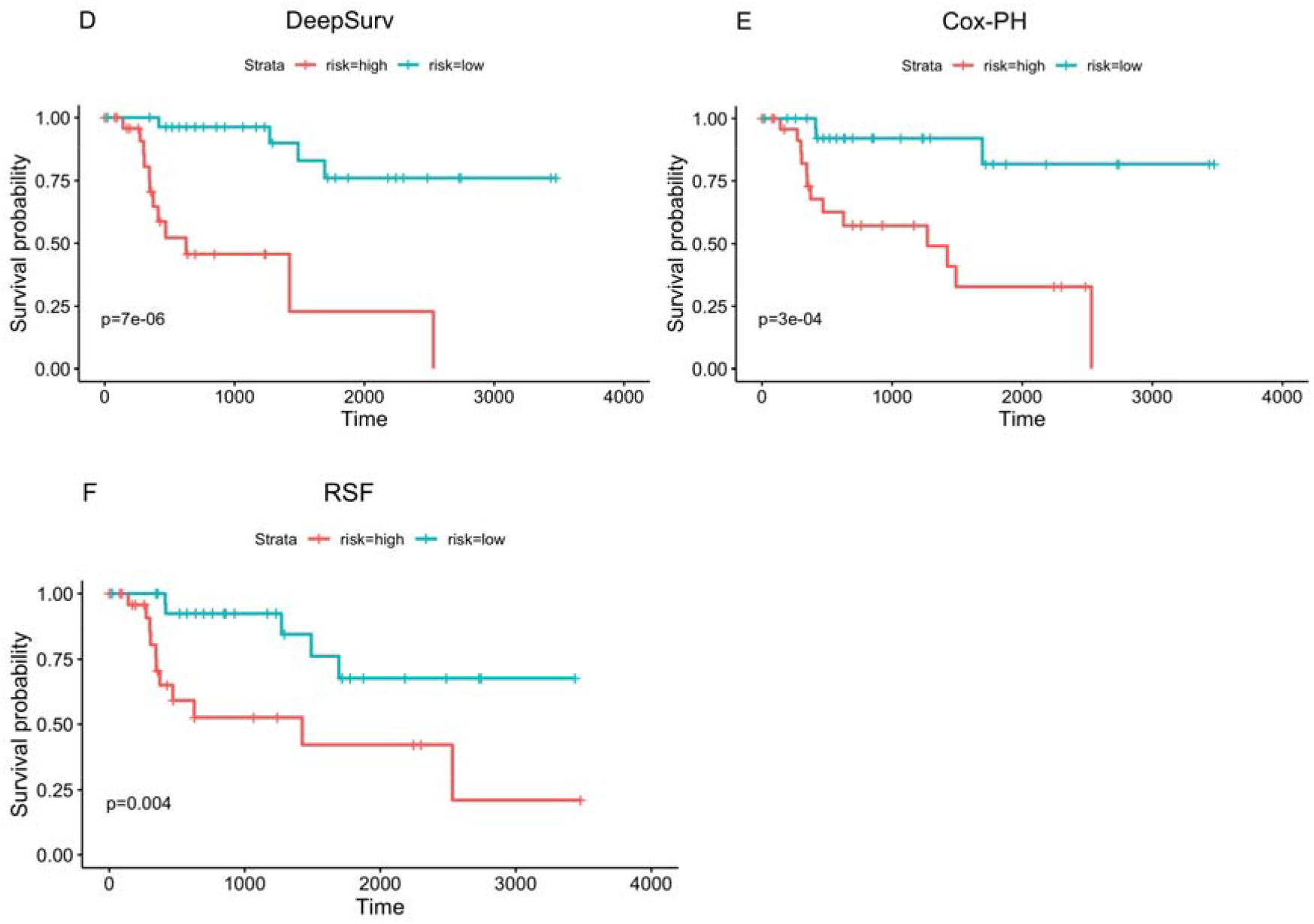
Comparison of prognosis prediction among different models using pathology imaging data. (A). C-index results on training and testing datasets. (B-F). Kaplan-Meier survival curves on testing datasets using different methods. (B) Cox-nnet (C) CoxBoost (D) DeepSurv (E) Cox-PH (F) Random Survival Forests (RSF).

### Two-stage Cox-nnet model to predict prognosis on combined histopathology imaging and gene expression RNA-Seq data

Multi-modal and multi-type data integration is challenging, particularly for survival prediction. We next ask if we can utilize Cox-nnet framework for such purpose, exemplified by pathology imaging and gene expression RNA-Seq based survival prediction. Towards this, we propose a two-stage Cox-nnet complex model, inspired by other two-stage models in genomics fields (23-25). The two-stage Cox-nnet model is depicted in **Figure 3**. For the first stage, we construct two Cox-nnet models in parallel, using the image data and gene expression data of HCC, respectively. For each model, we optimize the hyper-parameters using grid search under 5-fold cross-validation. Then we extract and combine the nodes of the hidden layer from each Cox-nnet model as the new input features for the second-stage Cox-nnet model. We construct and evaluate the second-stage Cox-nnet model with the same parameter-optimisation strategy as in the first-stage.

**Figure 3:**
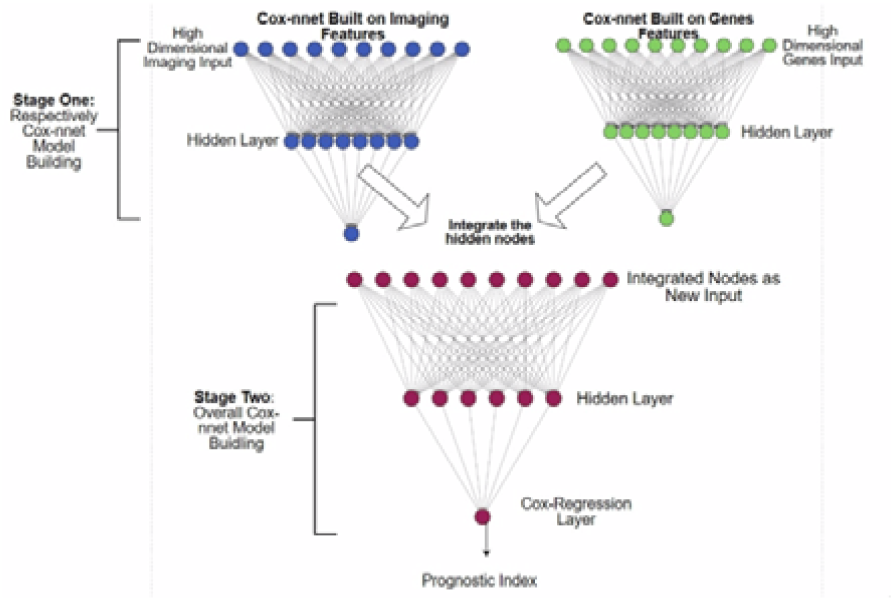
The architectures of two-stage Cox-nnet. The first stage builds individual Cox-nnet models for each data type. The second stage combines the hidden nodes from the first stage Cox-nnet models as the input, and builds a new Cox-nnet model.

As shown in **Figure 4A**, the resulting two-stage Cox-nnet model yields very good performance. Judging by the C-index values The median C-index scores for the training and testing sets are 0.89 and 0.75, respectively. These C-index values are significantly improved, compared to the Cox-nnet models that are built on either imaging (described earlier) or gene expression RNA-Seq data alone. For example, on the testing datasets, the median C-index score from two-stage Cox-nnet (0.75) is significantly higher (p<0.0005) than the Cox-nnet model built on gene expression data (0.70). It is also significantly higher (p<0.005) than the Cox-nnet model built on image data (0.74). The superior predictive performance of the two-stage Cox-nnet model is also confirmed by the log-rank p-values in the Kaplan-Meier survival curves (**Figure 4B-D**). It achieves a log-rank p-value of 6e-7 in testing data (**Figure 4D**), higher than the Cox-nnet models based on pathological image data (**Figure 4B**) or gene expression RNA-Seq data (**Figure 4C**), which have log-rank p-values of 4e-6 and 0.01, respectively.

**Figure 4:**
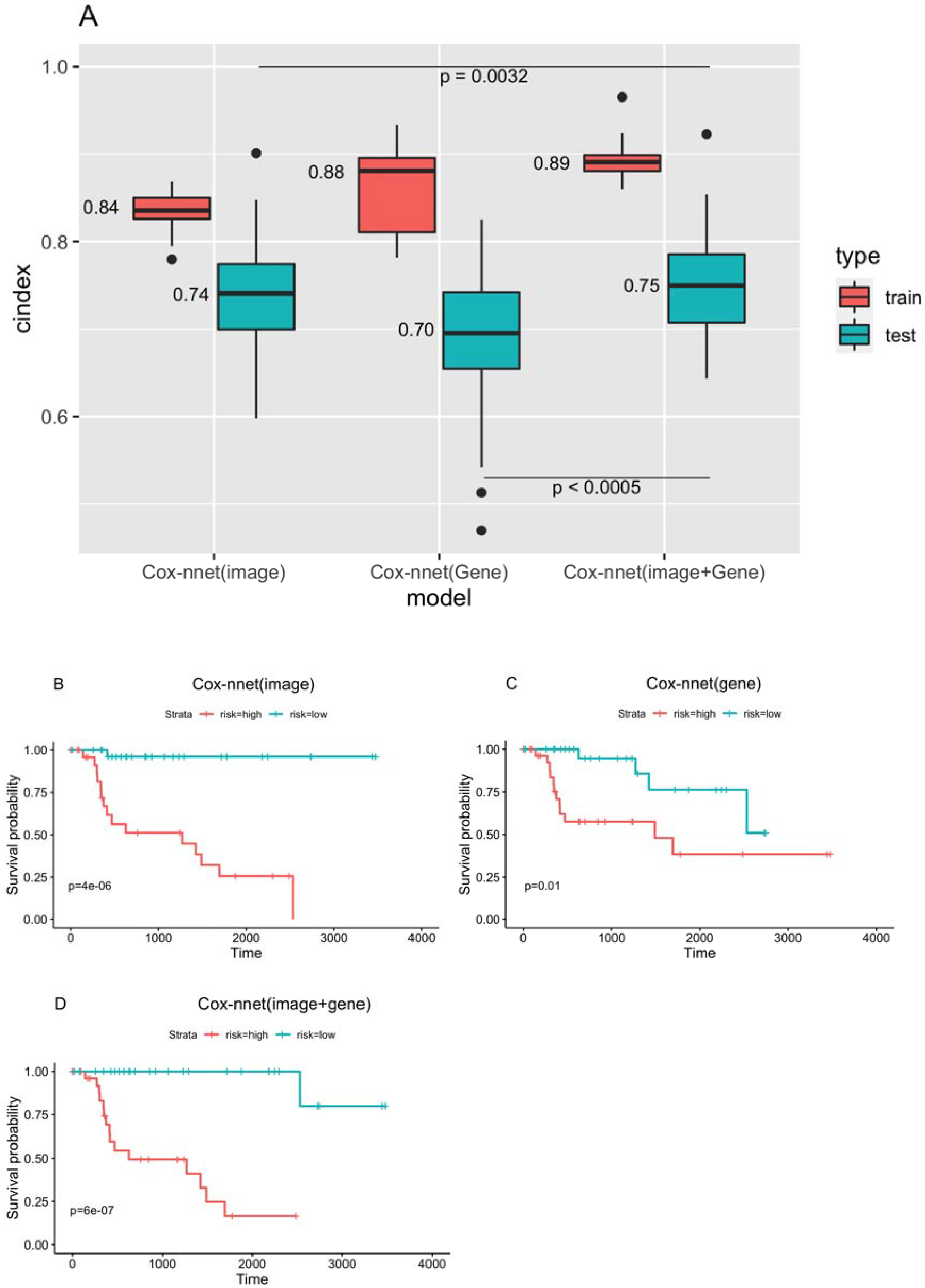
Comparison of Cox-nnet prognosis prediction using pathology imaging, gene expression and the combination of the two data types. (A). C-index results on training and testing datasets. (B-D). Kaplan-Meier survival curves on testing datasets. (B) Cox-nnet model on imaging data only. (C) Cox-nnet model on gene expression data only. (D) two-stage Cox-nnet model combining images and gene expression data.

### Comparing two-stage Cox-nnet model with other imaging and gene expression based prognosis models

We compare two-stage Cox-nnet with PAGE-Net, another neural-network method that combines imaging and genomics information to predict patient survival (5). The imaging prediction module of pagenet is very complex, with a patch-wide pre-trained convolutional neural network (CNN) layer followed by pooling them together for another neural network. The genomics model involves transformation of gene layer to pathway layer, and then followed by two hidden layers, the latter of which is combined with the image NN to predict patient survival. For a fair comparison, we use the same image and gene inputs for PAGE-Net and Cox-nnet. The data is randomly splitted into 80% training and 20% testing. The training set for PAGE-Net is further split into 90% training and 10% validation, which is used for early stopping. The whole experiment is repeated 20 times with different seeds.

As shown in **Figure 5A**, on the testing datasets, the median C-index score of 0.75 from the two-stage Cox-nnet model is significantly higher (p-value < 3.4e-4) than that of PAGE-Net (0.68). The C-index values from the PAGE-Net model are much more variable (less stable), compared to those from two-stage Cox-nnet. Moreover, PAGE-Net model appears to have an overfitting issue: the median C-index score of PAGE-Net model on the training set is very high (0.97), however, its predictability on hold-out testing data is much poorer. Moreover, impractical running time is another concern for PAGE-Net. Even on Graphic Processing Unit (GPUs) of Nvidia V100-PCIE with 16GB of memory each, it takes over a week to pretrain CNN and extract image features from only 290 samples, prohibiting its practical use. The Kaplan-Meier survival differences using median PI as the threshold confirm the observations by C-index (**Figure 5B-E)**. On testing data, two-stage Cox-nnet achieves a much better log-rank p-value of 6e-7 (**Figure 5D**), compared to that of 0.03 for PAGE-Net prediction (**Figure 5E**), even through Cox-nnet has a lower log-rank p-value of 4e-15 in training data (**Figure 5C**), compared to the value of 2e-30 of PAGE-Net (**Figure 5B**).

**Figure 5:**
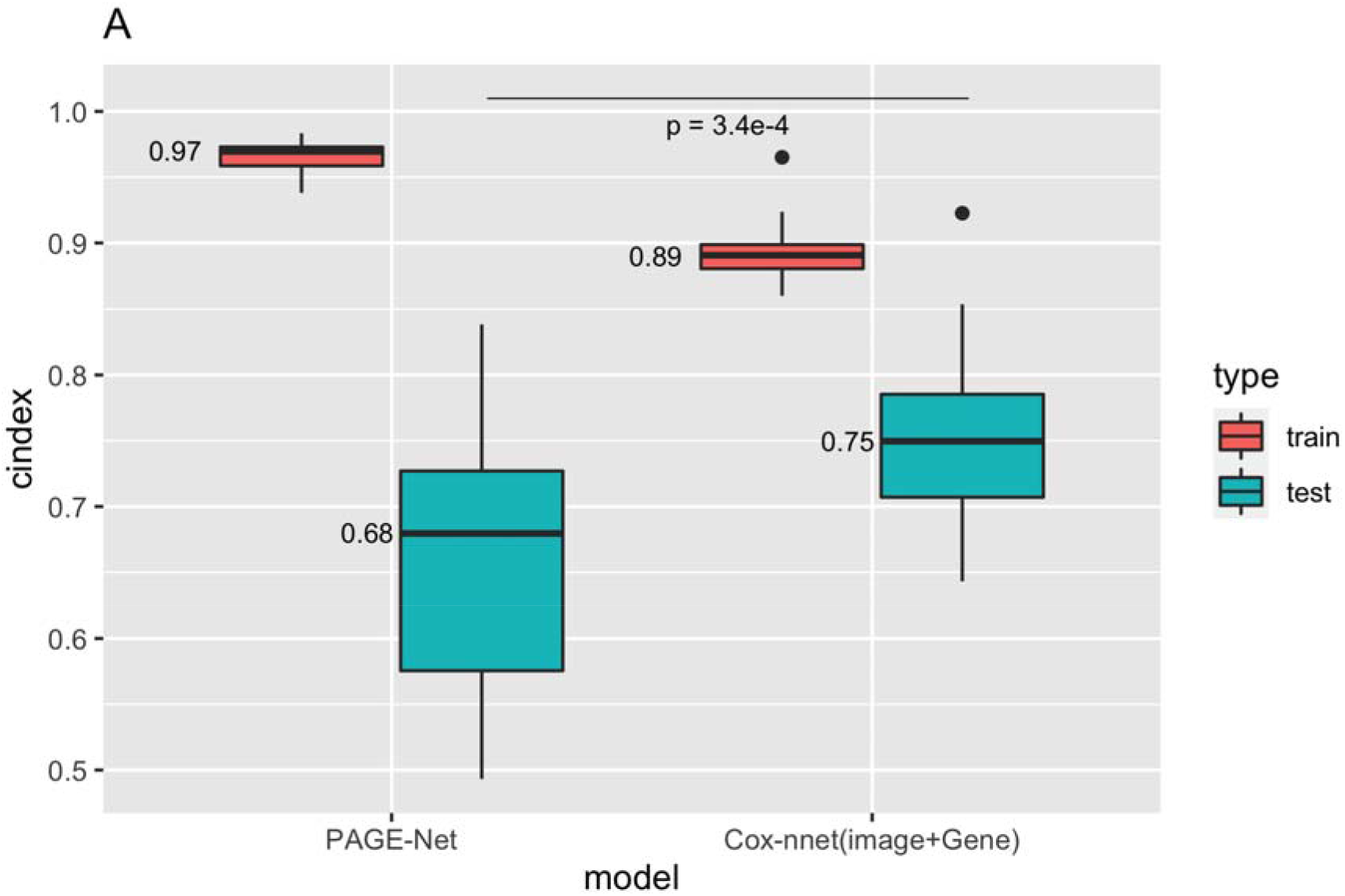
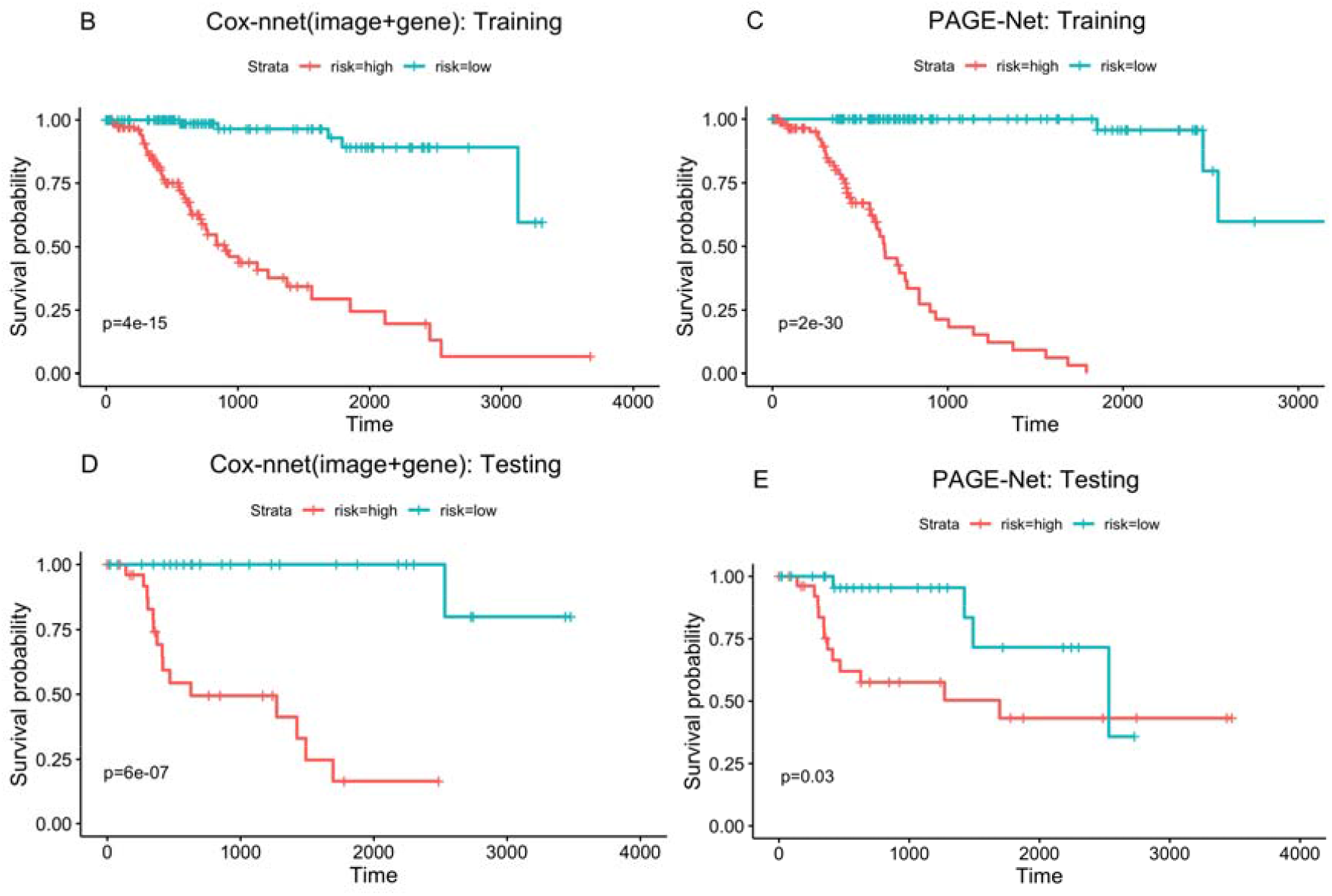
Comparison of two-stage Cox-nnet and PAGE-Net, based on combined pathological images and gene expression. (A) C-index of the two methods on training (red) and testing (blue) datasets, on 20 repetitions. (B-E) Kaplan-Meier survival curves resulting from the Cox-nnet (B, D) and PAGE-Net model (C, E) using training and testing datasets, respectively.

### Relationship between histology and gene expression features in the two-stage Cox-nnet model

We also investigate the correlations between the top imaging features and top RNA-Seq gene expression features. These top features are determined by the ranks of their feature scores (see Methods). The top 10 images and their Cox-nnet important scores are also listed in **Supplementary Table 1**. We then use the bipartite graph to show the image-gene pairs with Pearson’s correlations >0.1 and p-value<0.05, among the top 10 histopathology features and top 50 gene expression features (**Figure 6**). Top genes that are associated with top 10 histopathology features include long intergenic non-protein coding RNA 1554 (LINC01554), mucin 6 (MUC6), keratin 17 (KRT17), matrix metalloproteinase 7 (MMP7), secreted phosphoprotein 1 (SPP1) and myosin XVIIIB (MYO18B). Gene Set Enrichment (GSEA) analysis on top 1000 genes correlated to each image feature shows that image feature correlated genes are significantly (false discovery rate, FDR<25% as recommended) enriched in pathways-in-cancer (normalized enrichment score, NES=1.41, FDR=0.18) and focal-adhesion (NES=1.22, FDR=0.21) pathways, which are upregulated in HCC patients with poor prognosis. Pathways-in-cancer is a pan-pathway that covers multiple important cancer-related signalling pathways, such as PI3-AKT signaling, MAPK signaling and p53 signaling. Focal-adhesion pathway includes genes that involve cell-matrix adhesions, which play essential roles in important biological processes including cell motility, cell proliferation, cell differentiation, and cell survival. These two pathways play important roles in the survival of HCC patients (26).

**Figure 6:**
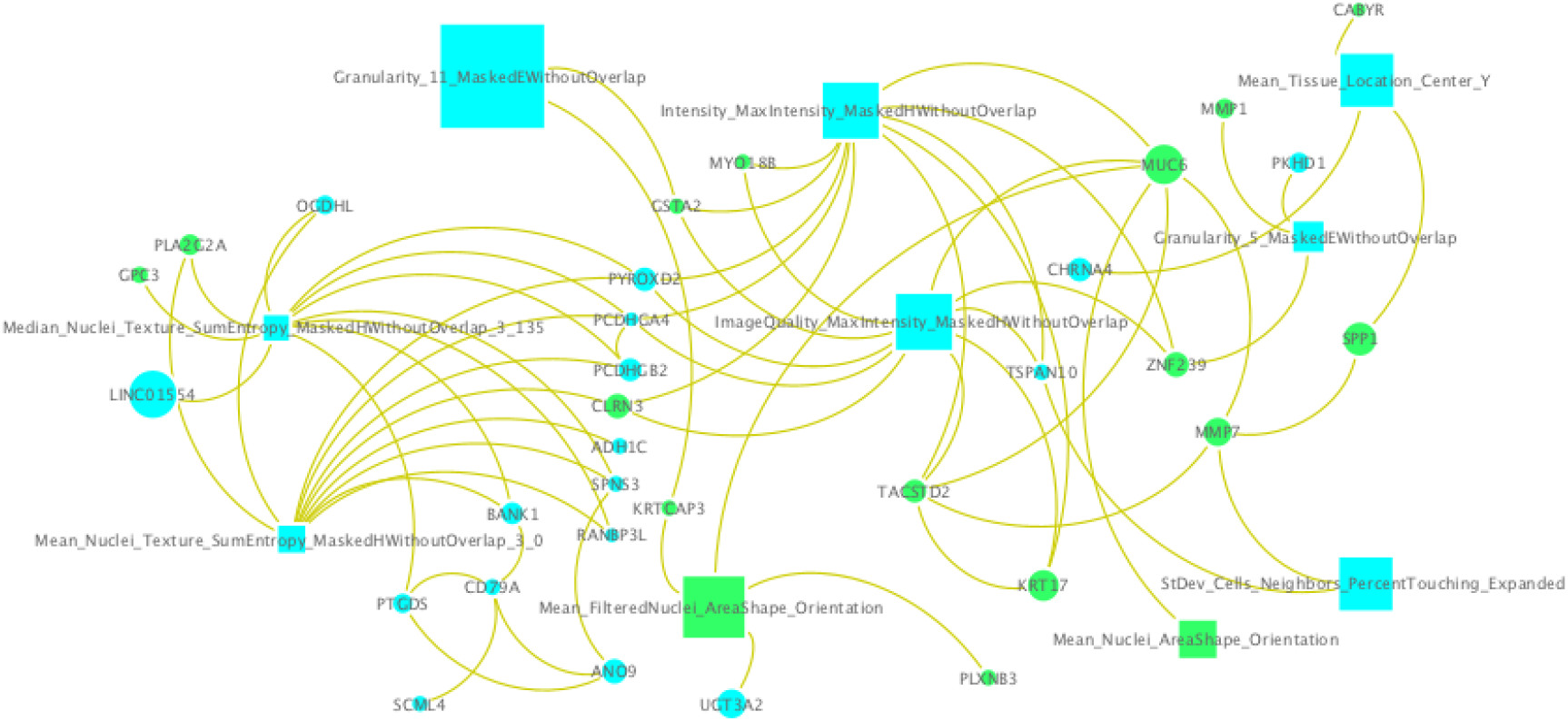
Relationship between top imaging and gene features. Rectangle nodes are image features and circle nodes are gene features. Node sizes are proportional to importance scores from Cox-nnet. Two gene nodes are connected only if their correlation is greater than 0.5; an image node and a gene node are connected only if their correlation is greater than 0.1 (p-value<0.05). Green nodes represent features with positive coefficients (hazard ratio) in univariate Cox-PH regression, indicating worse prognosis. Blue nodes represent features with negative coefficients (hazard ratio) in univariate Cox-PH regression, indicating protection against bad prognosis.

## 4 DISCUSSION

Driven by the objective to build a uniform workframe to integrate multi-modal and multi-type data to predict patient survival, we extend Cox-nnet model, a neural-network based survival prediction method, on pathology imaging and transcriptomics data. Using TCGA HCC pathology images as the example, we demonstrate that Cox-nnet is more robust and accurate at predicting survival, compared to Cox-PH the standard method which was also the second-best method in the original RNA-Seq transcriptomic study (1). We also demonstrate that Cox-nnet achieves better or comparable performance compared to other methods including DeepSurv, CoxBoost, and Random Survival Forests. Moreover, we propose a new two-stage complex Cox-nnet model to integrate imaging and RNA-Seq transcriptomic data, and showcase its superior accuracy on HCC patient survival prediction, compared to another neural network PAGE-Net. The two-stage Cox-nnet model combines the transformed, hidden node features from the first-stage of Cox-nnet models for imaging or gene expression RNA-Seq data respectively and uses these combined hidden features as the new inputs to train a second-stage Cox-nnet model.

Rather than using convolutional neural network (CNN) models that are more complex, such as PAGE-Net, we utilized a less complex but more biologically interpretable approach, where we extract imaging features defined by the tool *CellProfiler*. These features are then used as input nodes in relatively simple, two-layer neural network models. Hidden features extracted from each Cox-nnet model can then be combined flexibly to build new Cox-nnet models. On the other hand, PAGE-Net uses a pre-trained CNN for images and a gene-pathway layer to handle gene expression data. Despite great efforts, image features extracted by CNN in PAGE-Net are not easily interpretable, the model appears to be over-fit given the limited sample size, and requires very long training time. The significantly higher predictive performance of two stage Cox-nnet model argues for the advantages to use a relatively simple neural network model with input nodes of biological relevance, such as those extracted by imaging processing tools and gene expression input features.

Besides the interpretability of histopathology image features themselves, correlation analysis between top gene features and top image features identified genes known to be related to survival of HCC patients and/or morphology of the tissue, such as LINC01554, MUC6, MMP7, KRT17, MYO18B and SPP1. LINC01554 is a long non-coding RNA that is down-regulated in HCC and its expression corresponds to good survival of HCC patients previously (27). MUC6 is a mucin protein that participates in the remodeling of the ductal plate in the liver (28), which was also involved in the carcinogenesis of HCC (29). MMP7, also known as matrilysin, is an enzyme that breaks down extracellular matrix by degrading macromolecules including casein, type I, II, IV, and V gelatins, fibronectin, and proteoglycan (30). MMP7 participates in the remodeling of extracellular matrix (31) and impacts the morphology of liver tissue (32) which may explain its link to histopathology features. MMP7 expression is also associated with poor prognosis in patients with HCC (33). KRT17 serves as an oncogene and a predictor of poor survival in HCC patients (34). MYO18B is a myosin family gene that promotes HCC progression by activating PI3K/AKT/mTOR signaling pathway. Overexpression of MYO18B predicts poor survival of HCC patients (35). SPP1 functions as an enhancer of cell growth in HCC. The 5 year overall survival rate generated from 364 HCC cases demonstrated a poor survival of the patients with relatively higher SPP1 expression (36).

In summary, we extend the previous Cox-nnet model to process pathological imaging data, and propose a new class of two-stage Cox-nnet neural network model that creatively addresses the general challenge of multi-modal data integration, for patient survival prediction. Using input imaging features extracted from CellProfiler, Cox-nnet models are biologically interpretable. Some image features are also correlated with genes of known HCC relevance, enhancing their biologically interpretability. Since the proposed two-stage Cox-nnet is generic, we would like to extend this methodology to other types of cancers in the future, including those in TCGA consortium.

## Data Availability

All data used in this study is publicly available.

https://gs://gdc-tcga-phs000178-open/

## AUTHOR CONTRIBUTIONS

LXG envisioned the project, supervised the study and wrote the majority of the manuscript. ZZ and ZJ performed modeling and data analysis, with help from BH and NH. MW and EYC assisted with the tumor section labeling of all the images.

## ACKNOWLEDGEMENTS

We thank previous Garmire group members Dr. Fadhl Alakwaa, Dr. Olivier Poirion and Dr. Travers Ching for the discussions. LXG would like to thank the support by grants K01ES025434 awarded by NIEHS through funds provided by the trans-NIH Big Data to Knowledge (BD2K) initiative (www.bd2k.nih.gov), R01 LM012373 and R01 LM012907 awarded by NLM, and R01 HD084633 awarded by NICHD to L.X. Garmire.

