## Supplemental Table and Figures for "Two-stage biologically interpretable neural-network models for liver cancer prognosis prediction using histopathology and transcriptomic data"

**SUPPLEMENTARY MATERIALS**

**Supplementary Figure 1: Categories of the top 100 most important image features in Cox-nnet.**

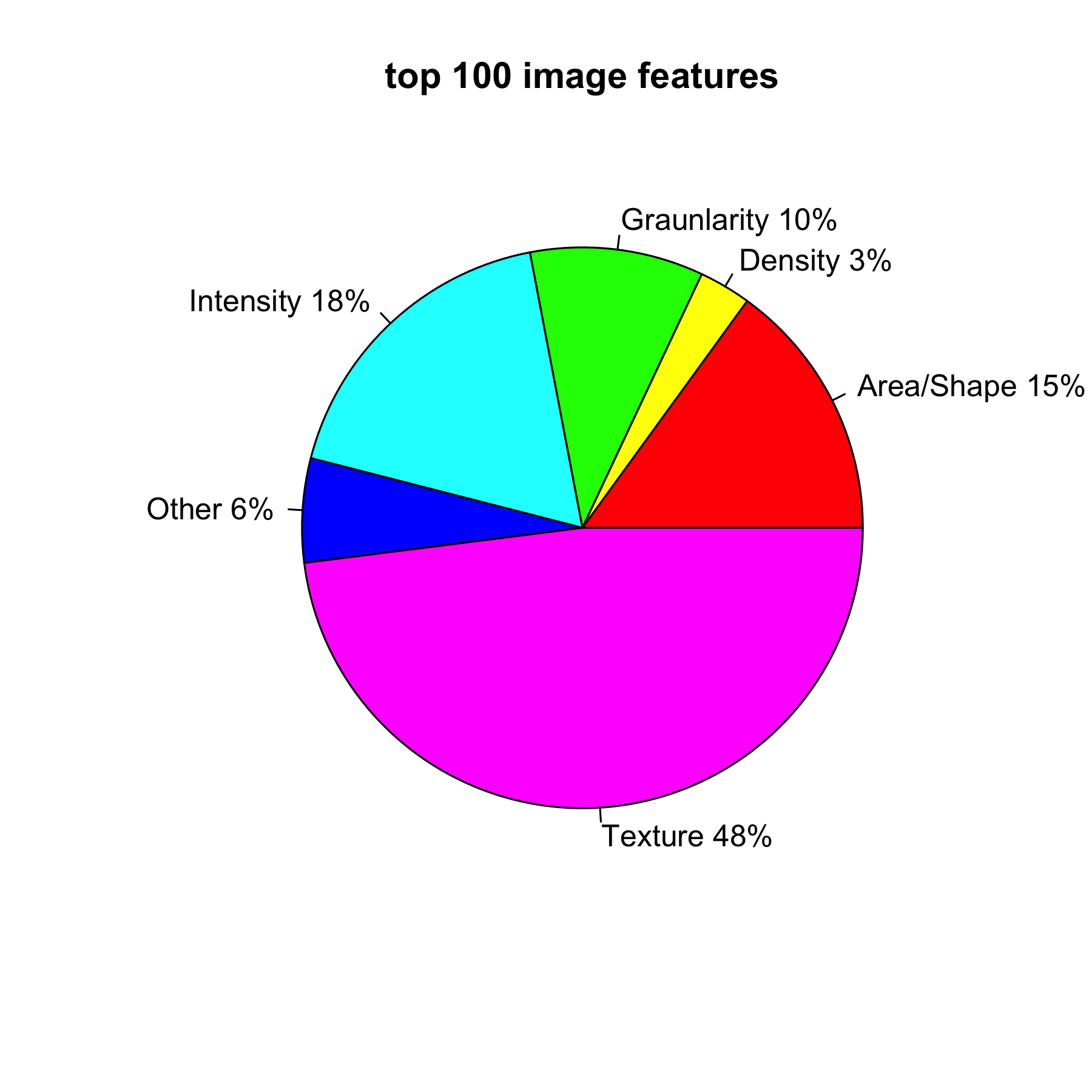

**Supplementary Figure 2: Train/valid loss curves for PAGE-Net and Cox-nnet models:**

(A) PAGE-Net	(B) Cox-nnet - image	(C) Cox-nnet - image+gene

The training loss curve of PAGE-Net fluctuates, maybe due to that the dataset has a small sample size (n=290) but high dimensionality (p=15,600), per communications with the authors of PAGE-Net.

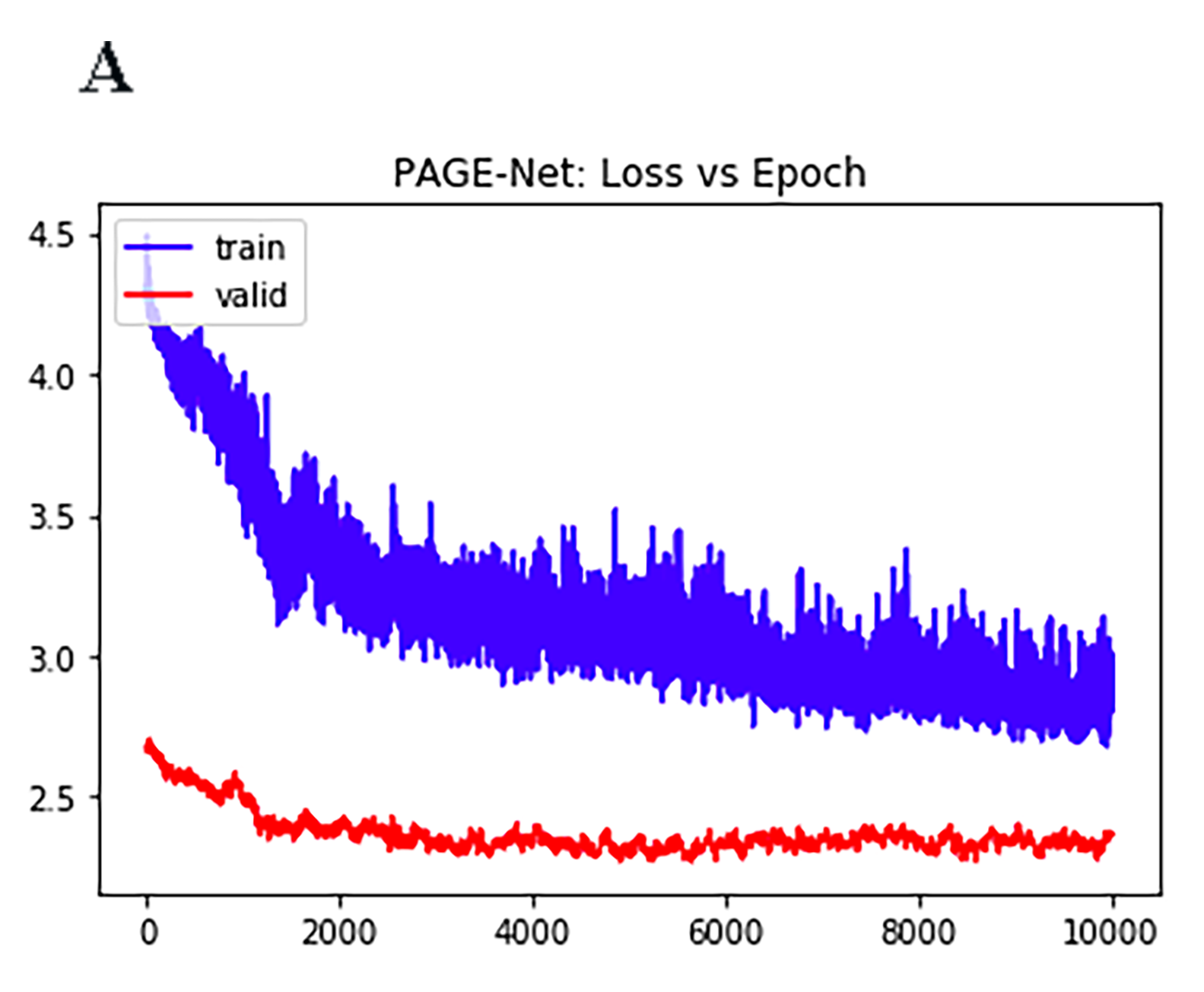

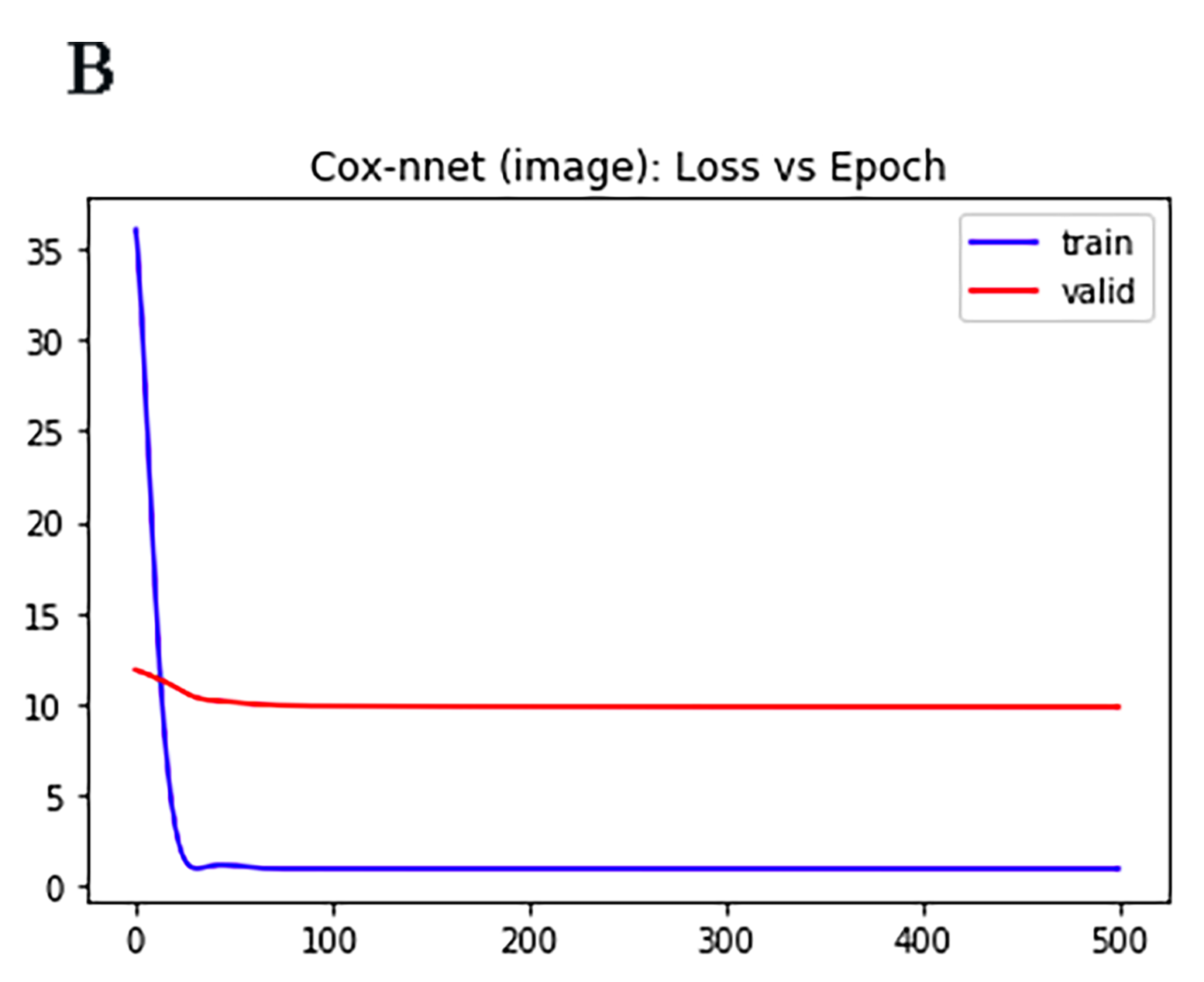

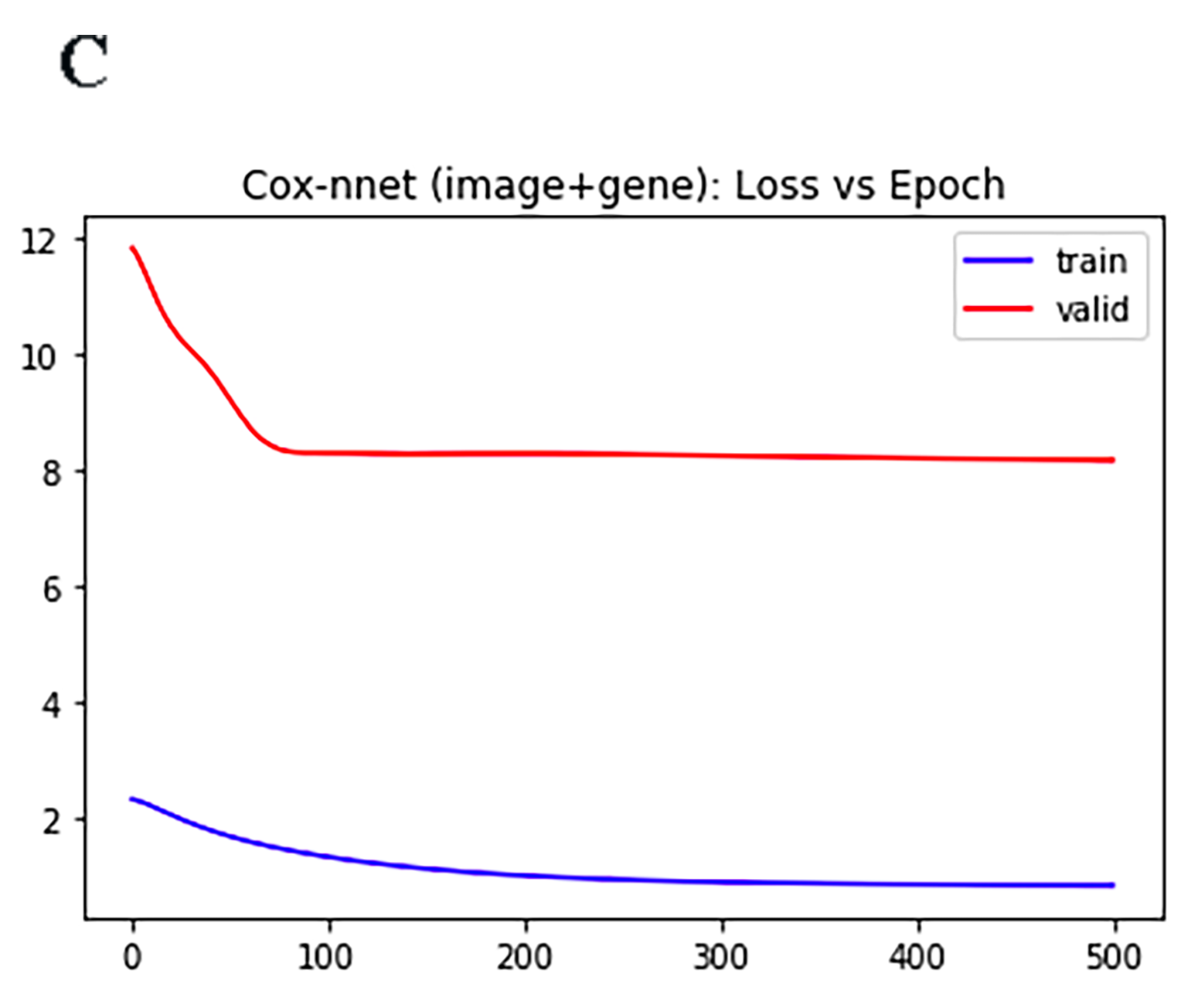

**Supplementary Figure 3: Comparison of prognosis prediction among different models using pathology imaging data.** Kaplan-Meier survival curves on training datasets using different methods. **(**A) Cox-nnet (B) CoxBoost (C) DeepSurv (D) Cox-PH (E) Random Survival Forests (RSF).

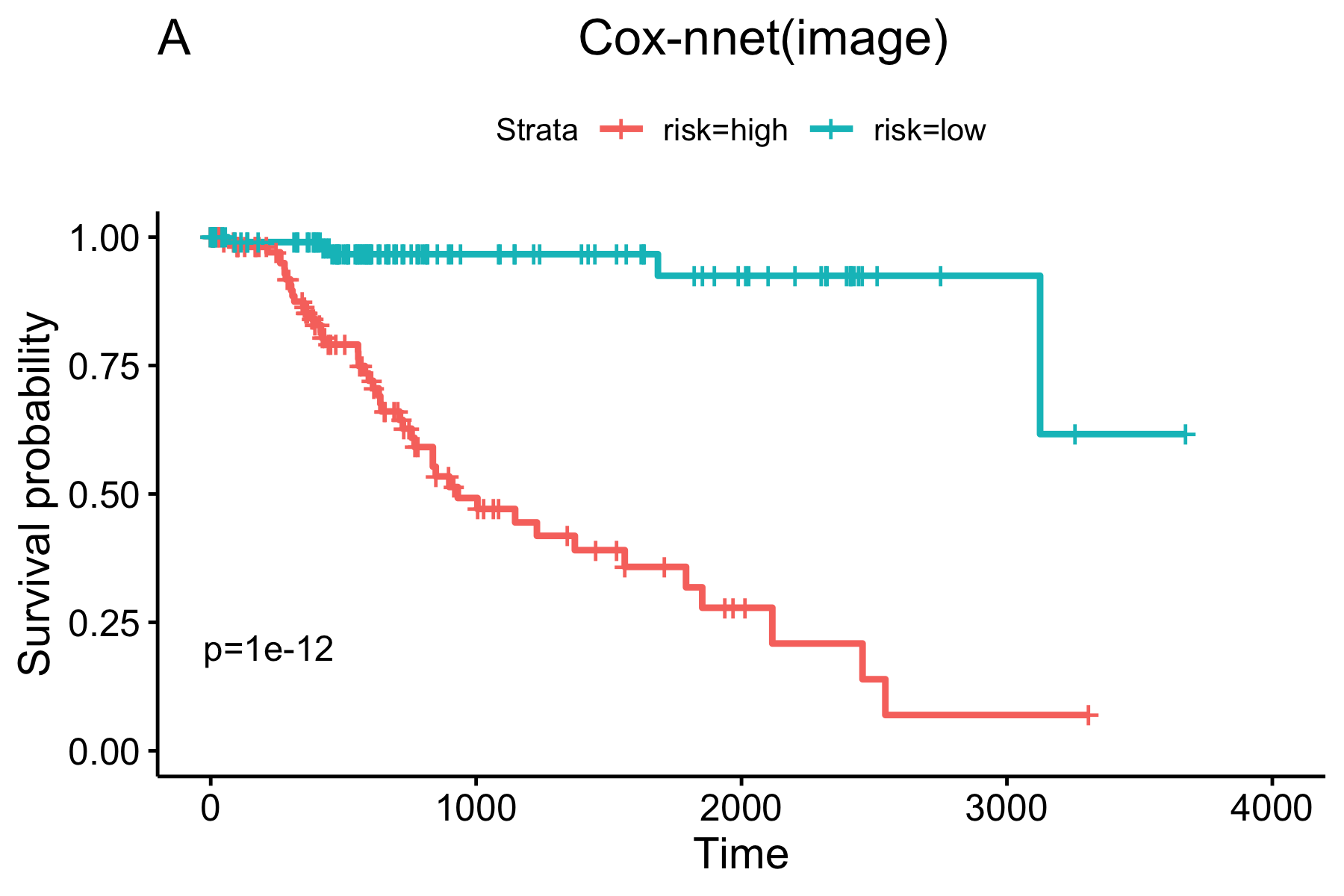

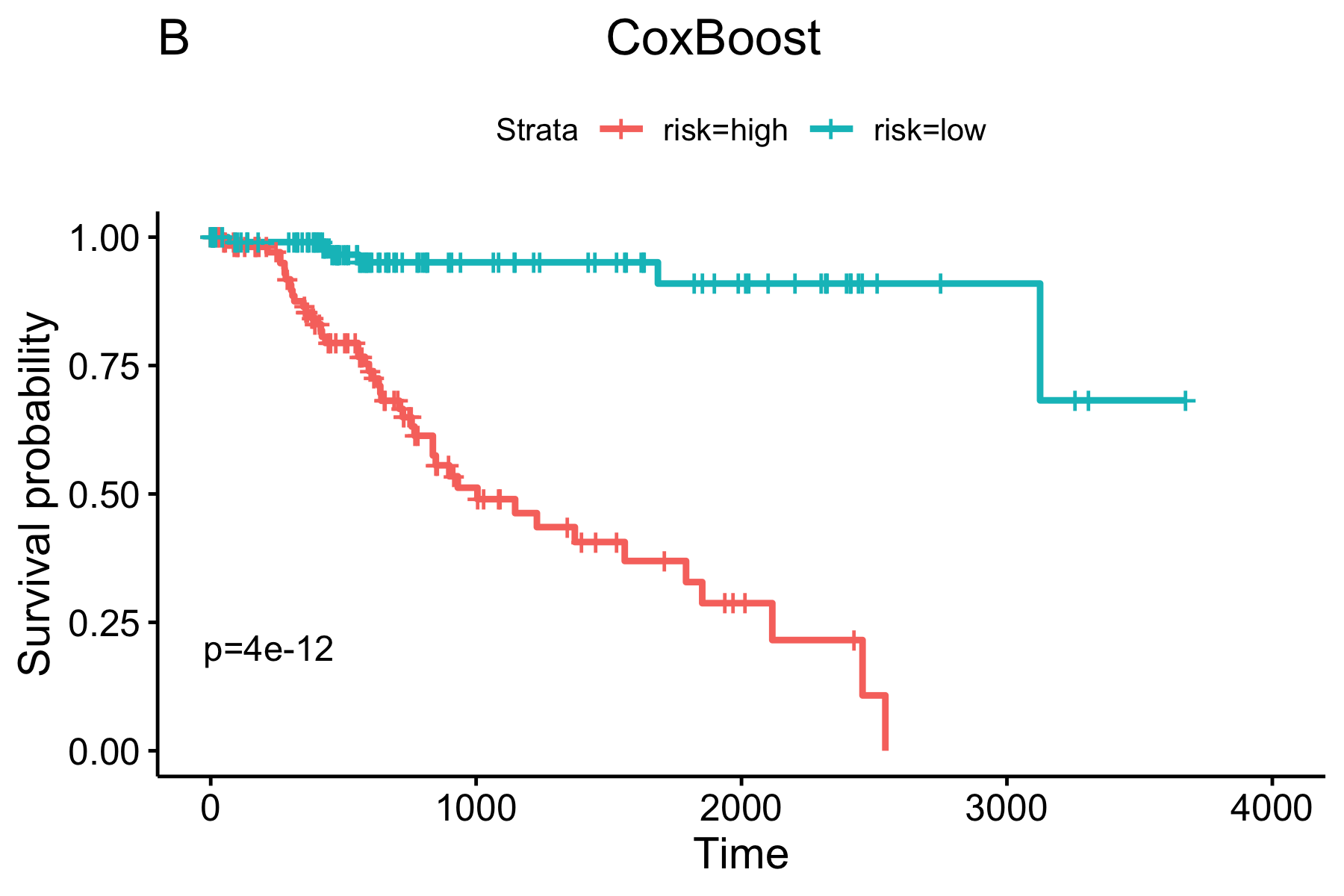

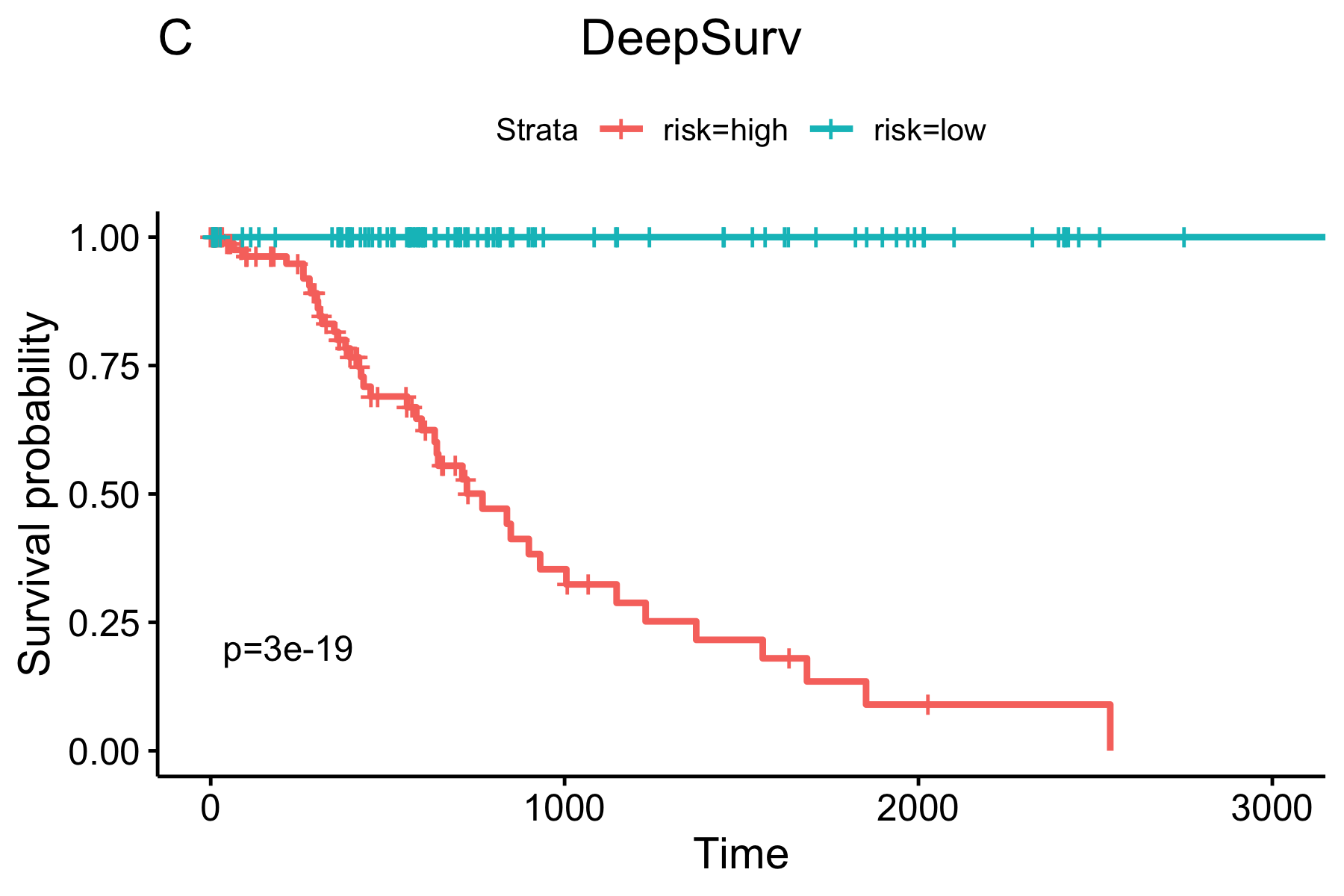

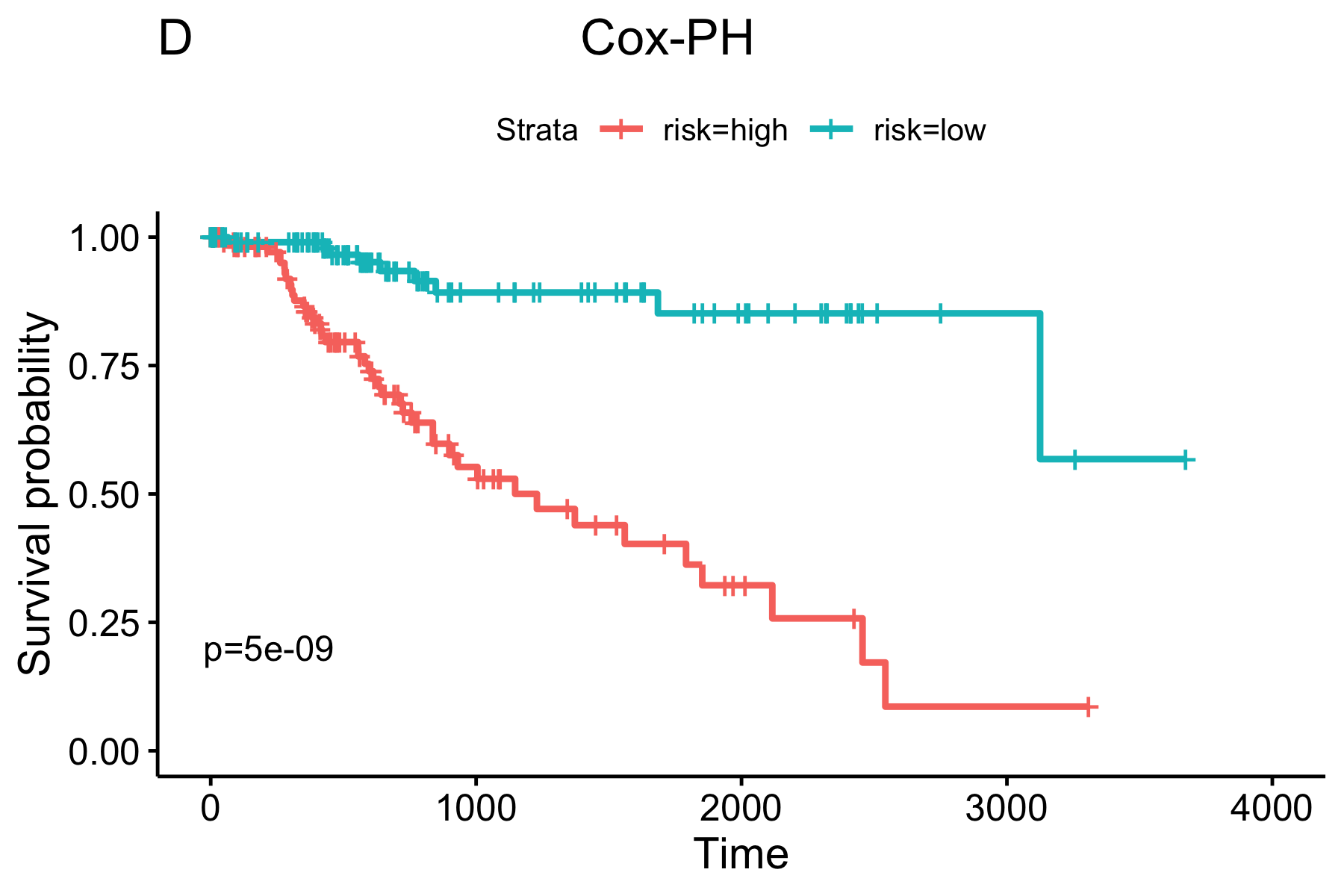

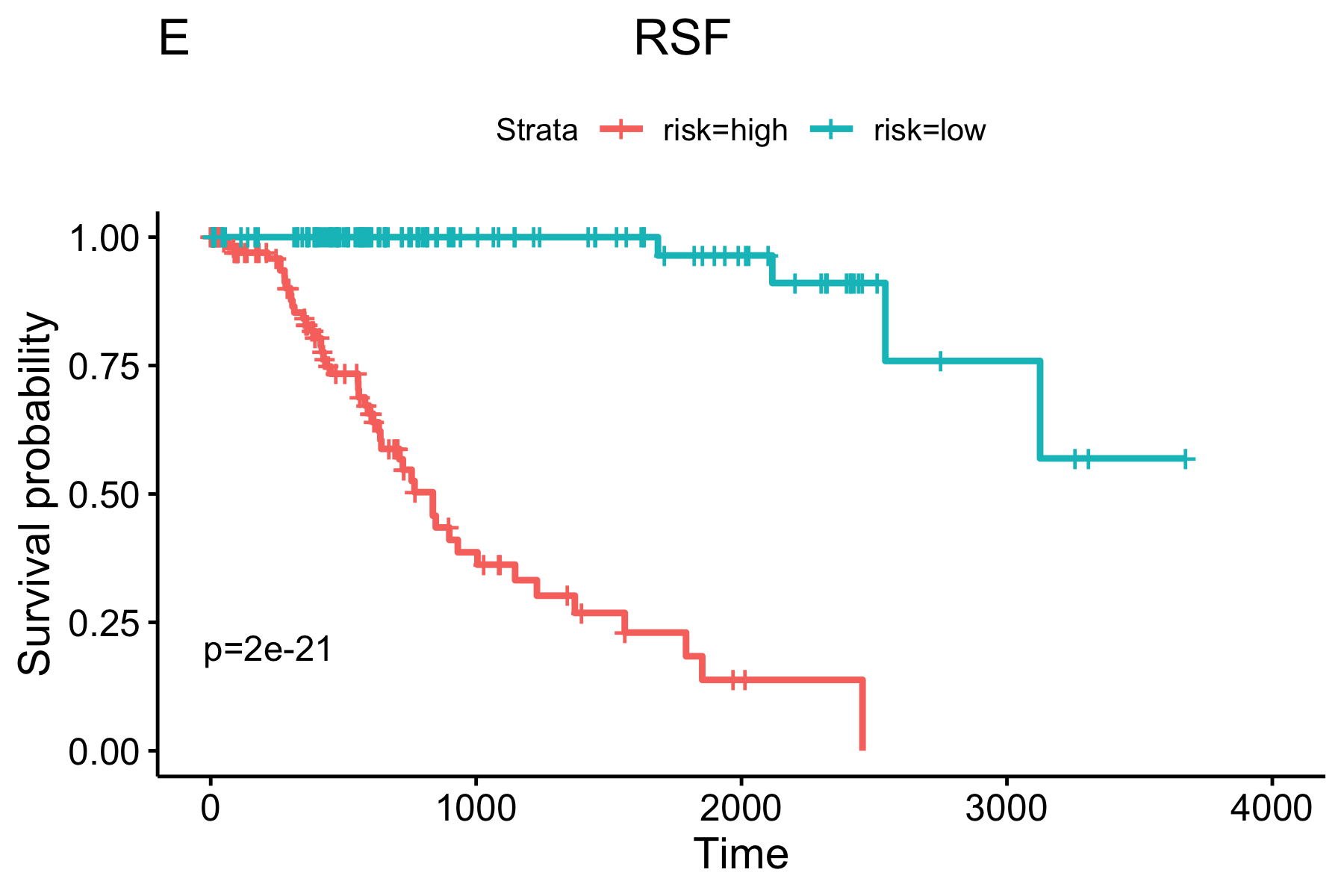

**Supplementary Table 1 :** Normalized feature importance scores of top 10 image features in Figure 6.

| **Top Image Feature** | **Importance score** |
| --- | --- |
| **Granularity_11_MaskedEWithoutOverlap** | **2.90** |
| **Mean_FilteredNuclei_AreaShape_Orientation** | **2.42** |
| **ImageQuality_MaxIntensity_MaskedHWithoutOverlap** | **2.34** |
| **Intensity_MaxIntensity_MaskedHWithoutOverlap** | **2.34** |
| **StDev_Cells_Neighbors_PercentTouching_Expanded** | **2.29** |
| **Mean_Tissue_Location_Center_Y** | **2.28** |
| **Mean_Nuclei_AreaShape_Orientation** | **1.99** |
| **Granularity_5_MaskedEWithoutOverlap** | **1.81** |
| **Mean_Nuclei_Texture_SumEntropy_MaskedHWithoutOverlap_3_0** | **1.72** |
| **Median_Nuclei_Texture_SumEntropy_MaskedHWithoutOverlap_3_135** | **1.67** |
